# Estimates of pandemic excess mortality in India based on civil registration data

**DOI:** 10.1101/2021.09.30.21264376

**Authors:** Murad Banaji, Aashish Gupta

## Abstract

**Background:** The population health impacts of the COVID-19 pandemic are less well understood in low-and middle-income countries, where mortality surveillance before the pandemic was patchy. Interpreting the limited all-cause mortality data available in India is challenging.

**Objective:** We use existing data on all-cause mortality from civil registration systems of twelve Indian states comprising around 60% of the national population to understand the scale and timing of excess deaths in India during the COVID-19 pandemic.

**Methods:** We carefully characterize the reasons why registration is incomplete and estimate the extent of coverage in the data. Comparing the pandemic period to 2019, we estimate excess mortality in twelve Indian states, and extrapolate our estimates to the rest of India. We explore sensitivity of the estimates to various assumptions.

**Results:** For the 12 states with available all-cause mortality data, we document an increase of 28% in deaths during April 2020–May 2021 relative to expectations from 2019. This level of increase in mortality, if it applies nationally, would imply 2.8–2.9 million excess deaths. More limited data from June 2021 increases national estimates of excess deaths during April 2020–June 2021 to 3.8 million. With more optimistic or pessimistic assumptions, excess deaths during this period could credibly lie between 2.8 million and 5.2 million. The scale of estimated excess deaths is broadly consistent with expectations based on seroprevalence data and international data on COVID-19 fatality rates. Moreover, the timing of excess deaths and recorded COVID-19 deaths is similar – they rise and fall at the same time.

**Conclusions:** The surveillance of pandemic mortality in India has been extremely poor, with around 8–10 times as many excess deaths as officially recorded COVID-19 deaths. India is among the countries most severely impacted by the pandemic. Our approach highlights the utility of all-cause mortality data, as well as the significant challenges in interpreting it.

## 1. Introduction

The COVID-19 pandemic has had large impacts on population health across the world. These impacts are less well understood in low- and middle-income countries (1–4), where routine mortality surveillance before the pandemic was patchy (5–7). Daily case counts and confirmed COVID-19 deaths have been widely used in policy and public discussions, including in India. These disease surveillance systems are innovative in the context of India. However, it is widely recognized that reported cases generally capture a small fraction of total infections, and that deaths in these systems are undercounts (8–10). All-cause mortality data, where available, has been used to understand the overall mortality impact of the pandemic (11,12). In India, numerous media reports have reported large increases in registered deaths (13–15). Owing to a variety of factors, interpreting these data and estimating the scale of pandemic excess deaths in India is challenging, however.

First, the data are available only for some of India’s 36 states and union territories. Second, vital registration varies substantially within the states from which all-cause mortality data are available (16–18). States differ in terms of level of mortality registration before the pandemic; recording systems; (for instance, for online and offline registration); and the way mortality data are organized (e.g., by date of death or date of registration) (19). Available data may miss not only the deaths that were not registered, but also sometimes deaths that were registered in an offline system (13). Third, levels and trends in registration and mortality need to be understood. There is uncertainty about baseline levels of mortality and registration completion (17,20–22). Often, registration was slowly increasing prior to the COVID-19 pandemic (6,23), but was disrupted during the early phase of the pandemic. This can be observed in data from the second phase of the fifth round of the National Family Health Survey (21). In the states visited by the NFHS-5 in 2021, deaths in 2020 were less likely to be registered than deaths that occurred in 2019 and 2018 (24). It is likely that some of this disruption was a consequence of national lockdown. It is not clear to what extent registration recovered as lockdown eased.

Carefully considering these concerns, we use reported all-cause mortality data from India’s civil registration system (CRS) to understand the scale and evolution of excess mortality in India during the COVID-19 pandemic. To do so, we compile contextual information on registration, baseline mortality, the extent to which available death records are complete, as well as trends and disruptions in death registration. We transparently lay out the many assumptions needed to estimate excess deaths on the basis of these data. Our approach draws on, and contributes to a large demographic and epidemiological literature on data quality of demographic indicators and estimates in data constrained settings (25–27). Given our careful attention to death records, these findings contribute to ongoing efforts to understand and improve civil registration systems. Because of India’s large population, epidemiological importance in the context of the COVID-19 pandemic, as well as vulnerability to mortality crises, our findings contribute to the global literature on the mortality impact of pandemics in low and middle income countries. Ultimately, our approach lays out plausible estimates of excess mortality in India given the data, paying attention to the quality of the data, the direction of biases, timing of deaths, and the sensitivity of estimates to different assumptions.

Our overall estimates are similar to those from complementary approaches (2,28–32). Our approach applies a careful adjustment process, accounting for both registration completion and coverage in online systems. We also pay close attention to pre-pandemic registration coverage, trends in registration, registration disruption during the 2020 lockdown in India, and the extent to which available registration data is complete. We use data from the twelve states from which monthly mortality records from January 2018 until at least May 2021 are publicly available. These states comprise around 60% of the national population. Moving beyond the existing literature, we consider the association in the timing of recorded COVID-19 deaths and excess deaths. We also assess the degree to which our estimates are sensitive to the various uncertainties, for example due to fluctuations in levels of registration, or possible differences in the mortality impact of the pandemic in the states for which we have data and the rest of the country.

Our findings indicate that the mortality impact of the pandemic in India has been severe. We estimate that the twelve states whose data we use saw around 28% more deaths than expected from historical data between April 2020 and May 2021. The limited data from June 2021 shows considerable further rises, perhaps partly as a result of delays in registration.

Our estimates of excess deaths are broadly consistent with expectations based on COVID-19 fatality rates given India’s age structure and the levels of spread estimated in seroprevalence surveys. On the other hand, by June 2021, we estimate a ratio of excess deaths to official COVID-19 deaths of over 9, indicating that official death counts have underestimated the scale of pandemic mortality by an order of magnitude.

Nationally, we estimate around 34% more deaths over a 15 month period from April 2020–June 2021 than expected from historical data. Uncertainties and incompleteness of the data, most importantly the possibility of changes in levels of death registration during the pandemic, mean that this surge could plausibly lie between 24% and 45%. These estimates place India amongst the harder hit countries in the world during the pandemic (33,34). In absolute terms, our central estimate amounts to around 3.8M excess deaths during April 2020–June 2021 with optimistic and pessimistic estimates of 2.8M and 5.2M excess deaths respectively. Although data from many countries is limited, consistent with other sources (2,32), these estimates make it likely that India is the country with the highest number of pandemic excess deaths in the world.

## 2. Methods and materials

### 2.1 The context of mortality and civil registration in India

To use registered deaths to estimate excess deaths in any given region, we need estimates of registration completion, namely the fraction of deaths which are registered, both before and during the pandemic. However, there are uncertainties around registration completion prior to the pandemic, and trends in completion.

Government estimates of completion rely on comparing registered deaths with expected deaths. The latter are derived using population estimates and survey-based estimates of the crude death rate (CDR). According to the 2018 Sample Registration System annual statistical report (35), henceforth, “the 2018 SRS report”, India’s CDR stood at 6.2 per 1K in 2018. Based on this estimate, registration completion in 2019 stood at 92%, as reported in the 2019 report on Vital Statistics of India based on the Civil Registration System (16), henceforth, the “2019 CRS report”. Estimates of completion based on the 2019 SRS report (36) will be even higher. There are, however, several reasons to believe that the estimated CDR of 6.2 is too low, and that completion in 2019 was less than 92% (17).

A variety of data sources and approaches detailed in Appendix 1, including from the UN population division (37) and the National Family Health Survey-5 (21), give estimates of the national CDR in 2019 ranging from 6.0 to 7.5 per 1K, corresponding to registration completion in 2019 from 76% to 96%. A number of calculations lead to estimates of death registration completion of around 80% (17,20,22). Most relevant from our point of view here, are sub-national estimates of registration completion in the 2019 CRS report based on sub-national estimates of CDR. From these sub-national estimates, a national CDR in 2019 of 6.6 per 1K can be derived, implying completion of around 86%. We use these sub-national estimates. We note that the estimates of completion may be somewhat too high in some states. This makes our estimates of pandemic excess mortality conservative.

While there is uncertainty around the national CDR prior to the pandemic, there is also uncertainty about how it was changing from year to year. According to the SRS bulletins (38), the national CDR saw a steady decline of around 1.5% per year from 6.5 in 2015 to 6.2 in 2018. In 2019 and 2020, estimates of CDR dropped further to 6.0 deaths per 1,000. According to UN estimates (39), however, India’s estimated CDR was falling prior to 2015 and but saw a 1% increase during 2015–2019. Meanwhile, population projections suggest that the national population has been growing by around 1% per year (40). This is also the estimated population growth rate in states whose data we use below. Thus, the SRS estimates imply that up to 2019 year-on-year deaths were steady or falling slightly; while UN estimates suggest they could have been rising by a little over 1% per year.

While the estimated changes in yearly deaths are fairly small, there was a larger increase in estimated registration completion between 2018 and 2019. According to the 2019 CRS report (16), completion nationally increased from 84.6% to 92% between 2018 and 2019. Using the sub-national data in this report, we find an increase in estimated registration completion from 81.1% to 86.5% nationally. Completion increased from 86.9% to 92.1% between 2018 and 2019 in the twelve states from where we use data. Incomplete but increasing death registration in Bihar was a key factor in this increase: if we remove Bihar from the picture, the remaining eleven states together saw estimated registration completion rise from 95.7% to 98.4% between 2018 and 2019. All of these estimates assume no change in national or sub-national values of the CDR between 2018 and 2019. It is worth noting that according to these estimates registration was approaching 100% in many states.

### 2.2 The available data from twelve states

Henceforth, unless stated otherwise, all estimates of registration completion during 2019 are based on the sub-national data from the 2019 CRS report, as discussed above.

We use civil registration data from the following twelve states to arrive at estimates of excess mortality in India: Andhra Pradesh, Bihar, Haryana, Himachal Pradesh, Karnataka, Kerala, Madhya Pradesh, Maharashtra, Punjab, Rajasthan, Tamil Nadu and West Bengal. We refer to these states collectively as STAR12 – a rough mnemonic for “12 States with Available Registration Statistics”. In these states, partial or complete death registration data are available for at least January 2018 to May 2021. Data are additionally available for June 2021 from Andhra Pradesh, Karnataka and Punjab. The full data and code used in our analysis are available on our replication repository on GitHub (41).

STAR12 accounted for 59% of the estimated 2019 national population and also 59% of estimated total deaths occurring during 2019 (40). The estimated CDR in STAR12 was thus close to the national value. On the other hand, registration completion in STAR12 was somewhat higher than the national average: around 92% of deaths occurring during 2019 in STAR12 were registered, as against 78% in the remainder of the country. These estimates are obtained by comparing registered deaths in the 2019 CRS report to total deaths estimated using subnational completion estimates in the same report. The calculations are available at our replication repository (41).

Often, publicly available data do not include all registered deaths because they are from online systems which may miss deaths registered using offline systems. The available data includes 4,102,882 deaths, or 85% of the 4,811,595 deaths registered in STAR12 during 2019. Deaths recorded in the available data amount to 79% of the 5,222,286 deaths estimated to have occurred in these states during 2019 according to the SRS. We reference this fact by saying that “coverage in the data” was 79% in 2019, i.e., the fraction of estimated total deaths in 2019 in STAR12 which appear in the data was 79%. A flowchart depicting the attrition from estimated deaths to registered deaths recorded in the available data is shown in Figure 1.

**Figure 1:**
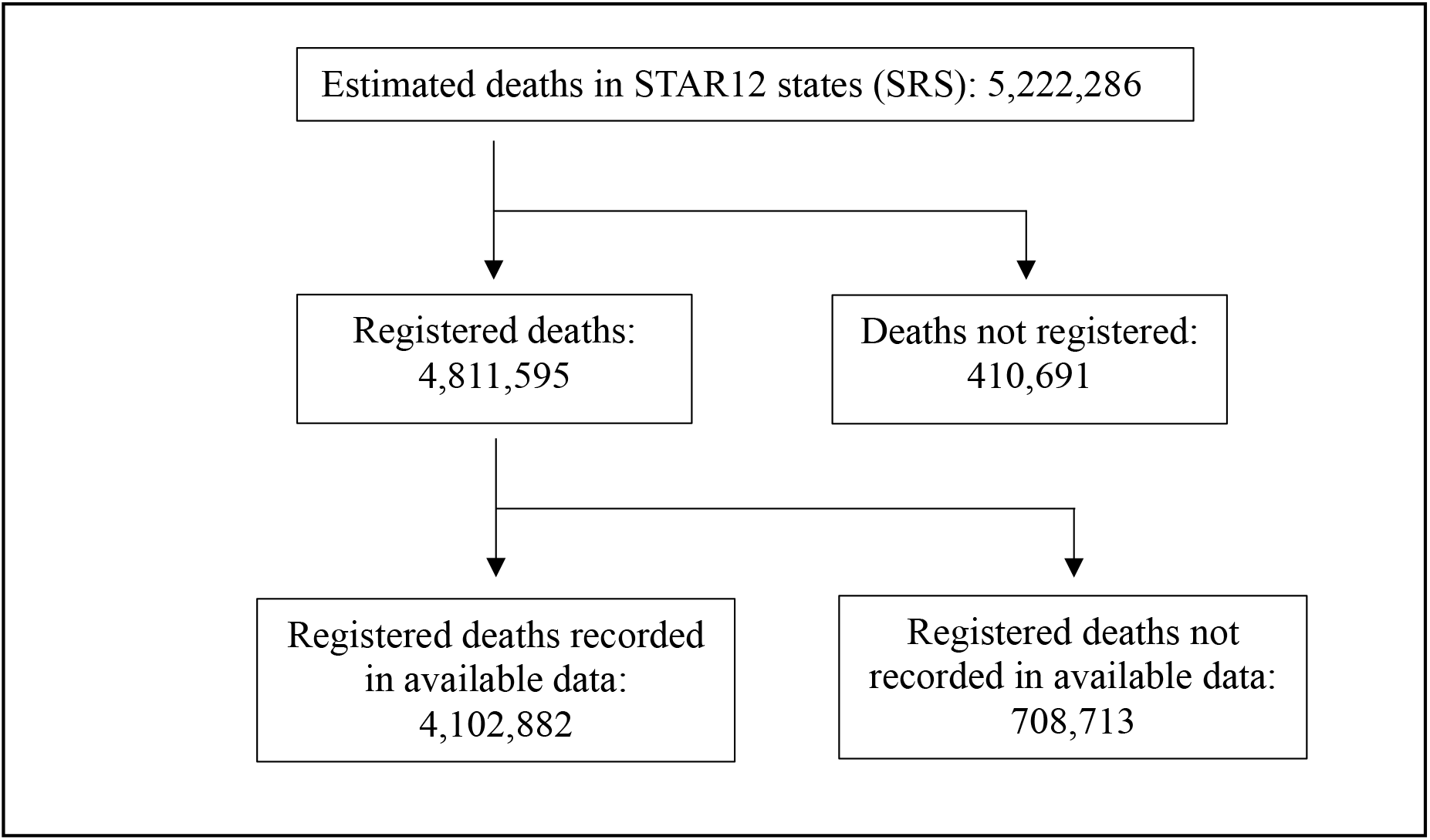
Coverage of estimated deaths for STAR12 states in 2019

Table 1 shows that in individual states in STAR12 in 2019, death registration varied between 51.6% and 100%. Coverage in the data during 2019 varied from 48% to 100%.

**Table 1:** Estimated deaths, registration, and coverage in STAR12, 2019.

| **Deaths, registration, and coverage in 2019** |  |  |  |  |  |
| --- | --- | --- | --- | --- | --- |
|  | **Estimated deaths** | **Registered deaths** | **Estimated registration completion (%)** | **Deaths recorded in the available data** | **Estimated coverage in the available data (%)** |
| **Andhra Pradesh** | 401,472 | 401,472 | 100.0 | 363,649 | 90.6 |
| **Bihar** | 696,413 | 359,349 | 51.6 | 351,248 | 50.4 |
| **Haryana** | 188,910 | 188,910 | 100.0 | 183,795 | 97.3 |
| **Himachal Pradesh** | 50,501 | 43,633 | 86.4 | 40,970 | 81.1 |
| **Karnataka** | 508,584 | 508,584 | 100.0 | 508,584 | 100 |
| **Kerala** | 270,567 | 270,567 | 100.0 | 264,071 | 97.6 |
| **Madhya Pradesh** | 553,679 | 493,328 | 89.1 | 449,819 | 81.2 |
| **Maharashtra** | 693,800 | 693,800 | 100.0 | 462,028 | 66.6 |
| **Punjab** | 215,045 | 215,045 | 100.0 | 213,122 | 99.1 |
| **Rajasthan** | 457,723 | 451,315 | 98.6 | 219,814 | 48.0 |
| **Tamil Nadu** | 633,897 | 633,897 | 100.0 | 588,221 | 92.8 |
| **West Bengal** | 551,695 | 551,695 | 100.0 | 457,561 | 82.9 |
| **Total (STAR12)** | **5,222,286** | **4,811,595** | **92.1** | **4,102,882** | **78.6** |

**Note:** Registered deaths and estimates of completion from the 2019 CRS report are used to estimate total deaths during 2019 in STAR12 states. These can be compared to the number of deaths in the available data to get estimates of coverage in the data during 2019.

Coverage in the data in STAR12 rose from 73% in 2018 to 79% in 2019. Most of this increase reflects an increase in registration completion rather than, say, increasing use of online systems: in fact, the fraction of *registered* deaths appearing in the available data rose only slightly, from 83.6% in 2018 to 85.3% in 2019. If we were to omit Bihar, the data appears even more stable: the remaining eleven states saw coverage in the data rise only modestly from 80% in 2018 to 83% in 2019.

The recently released 2020 CRS report (42) does not provide monthly registration data; however, total death registrations for 2020 given in the report for states in STAR12 are broadly consistent with estimates from the monthly data used in this study (24).

### 2.3 Methods

Using historical data to predict expected deaths, key to estimating excess deaths, is challenging in the Indian context. Pre-pandemic trends in registered deaths mainly reflect trends in registration completion rather than in mortality. On the other hand, with registration at or close to completeness in many states (according to SRS estimates), and with considerable disruption to registration during the pandemic, it could be misleading to extrapolate past trends of increasing registration completion into the pandemic period. For this reason, we choose a relatively simple approach. For our central estimates, expected deaths are set at estimated 2019 levels; but we explore how changing levels of registration would affect the resulting estimates of excess mortality. We see this approach as sufficient for the purposes of making aggregate estimates; but making credible state-level estimates would require a more careful case-by-case analysis. Preliminary state-factsheets which attempt such state-level analysis are available at India COVID Mapping (19). This analysis indicates that pre-pandemic trends in registration, and the extent to which these trends were disrupted by the pandemic, may have differed in different states.

We proceed as follows. In each state for which the data is available, we set baseline expectations for death registrations in a given pandemic month to be registrations during the corresponding month in 2019. Monthly excess registrations, namely, the difference between observed and expected registrations each month, can be summed over any pandemic period to obtain excess registrations during this period. We then use state-level estimates of coverage in the data (see Table 1) to scale excess registrations in each state, obtaining estimates of excess deaths in that state.

Following this procedure, and summing over all the states in STAR12, we obtain estimates of excess deaths in STAR12 during April 2020–May 2021. We can then estimate excess deaths nationally using two approaches:

1. **Mortality-based extrapolation**. We assume that per capita pandemic excess deaths nationwide were equal to in STAR12. For the denominator in these calculations, we use estimated 2020 populations.
2. **P-score-based extrapolation**. We refer to the percentage increase in mortality during any given pandemic period – relative to expected baseline mortality – as the P-score (33,43). For P-score-based extrapolation, we assume that the P-score nationwide was similar to that in STAR12. As we do not have national monthly data from the pre-pandemic period, we take monthly deaths nationally during 2019 to be one twelfth of the estimated yearly total.

Using the limited data available for June 2021, the same approaches to extrapolation can be used to extend the national estimates of excess deaths to the period April 2020– June 2021.

We remark that other approaches to extrapolation are possible: for example, we might attempt to use recorded COVID-19 deaths as a basis for extrapolation, i.e., to assume the ratio of excess deaths to COVID-19 deaths seen in STAR12 holds nationally. We reject this approach for reasons discussed in Section 4 below.

We bear in mind that there may have been shifts in expected mortality, and in coverage in the data during the pandemic, and carry out extensive analysis of how these might affect the estimates later in robustness checks. These robustness checks help bound our central estimates. We follow two approaches to do this: a scenario-based approach, and a Monte-Carlo simulations-based approach.

Overall, we consider the effects on estimated excess mortality of four factors: pre-pandemic registration completion; changes in coverage in the data between 2019 and the pandemic period; changes in expected deaths between 2019 and the pandemic period; and possible differences in the mortality impact of the pandemic in STAR12 and the rest of the country. A basic sensitivity analysis in Appendix 2 reveals that of these four factors, it is changes in coverage in the data which can potentially cause the greatest errors in estimated excess mortality. Based on this analysis, we examine two scenarios which we consider plausible best- and worst-case scenarios. Table 2 summarises the scenarios:

**Table 2:** Optimistic and pessimistic scenarios to assess sensitivity of excess mortality estimates

| **Factors** | **Optimistic Scenario** | **Pessimistic Scenario** |
| --- | --- | --- |
| Change in expected deaths (associated with population growth and natural increases in CDR) | There was a 2% increase in expected deaths in each state. | There was no change in expected deaths relative to 2019 levels. |
| Baseline coverage | Baseline coverage in the data was correctly estimated. | Baseline coverage in the data was overestimated by 5% in each state. |
| Change in coverage in the data | There was a 5% relative increase in coverage in the data. In other words, in each state, the fraction of total deaths which appear in the available data increased by 5% during the pandemic period relative to 2019 levels; but we cap registration completion in any given state at 100%. | There was a 5% relative decrease in coverage in the data in each state during the pandemic period. |
| Mortality in other states | The remaining states and territories saw a 20% lower P-score/excess mortality than STAR12. | The remaining states and territories saw a 20% higher P-score/excess mortality than STAR12. |

In the simulation approach, we ran Monte-Carlo simulations allowing baseline coverage in the data to vary by ±15% around the midpoint value estimated from the 2019 CRS report, and pandemic period coverage in the data to change by ± 5% in each state relative to its baseline value. In all cases, we capped coverage at 100%, i.e., any values of coverage greater than 100% were set at 100%. We also allowed the mortality impact outside of STAR12 to vary between 80% and 120% of the impact in STAR12. Choosing uniform distributions on all parameters, and running 10,000 simulations allowed us to generate confidence intervals on the mortality estimates for the period April 2020–June 2021.

## 3 Results

### 3.1 Central estimates of excess mortality in STAR12 during April 2020–May 2021

We begin by considering the 14-month period April 2020–May 2021 for which we have data from all states in STAR12. We find 1.3M excess registrations in STAR12 during April 2020–May 2021, amounting to 27% more registrations than expected from 2019 data.

If we scale the excess registrations in each state based on estimated coverage in the data during 2019 (see Table 1), we find 1.7M excess deaths during April 2020–May 2021 in STAR12, corresponding to a P-score of 28% in these states relative to expectations from 2019. This amounts to 2.1 excess deaths per thousand population in STAR12 during April 2020–May 2021. These estimates assume there were no variations in expected mortality, or in coverage in the data between 2019 and the pandemic period.

### 3.2 National estimates of excess mortality in STAR12 during April 2020–May 2021

Extrapolating from observed excess mortality in the STAR12 states, we now estimate levels of excess mortality nationally. We can do this via mortality-based extrapolation or P-score-based extrapolation, as defined in section 2.3. The fact that the estimated crude death rate in STAR12 matches the national estimate (see section 2.2) means that these two approaches give similar results: 27–28% more deaths nationally during April 2020– May 2021 than expected from 2019 data. This amounts to 2.8–2.9M excess deaths in this period. We discuss later the effects on our estimates if the mortality impact in regions outside STAR12 was higher or lower than in STAR12.

### 3.3 Extending the national estimates to June 2021

Data for June 2021 is currently only available for three states of STAR12: Andhra Pradesh, Karnataka, and Punjab, which hold around 11% of the national population. During June 2021, registrations in these states were 123% higher than registrations during June 2019. Moreover, the ratio of excess deaths to recorded COVID-19 deaths in these three states jumped from 8.0 in May 2021 to 11.7 in June 2021. This suggests that delays in registration following the enormous mortality surge in May could be at least partly responsible for the high June excess deaths.

We can use either mortality-based or P-score-based extrapolations from these states to obtain national estimates of excess deaths for June 2021. When we add these to the estimates for April 2020–May 2021, we find estimates of national excess deaths during April 2020–June 2021 of 3.8M. This amounts to an increase of 34% over deaths expected during a 15-month period from 2019 data, or around 2.8 excess deaths per 1K. Clearly, these figures carry greater uncertainty, given the limited data from June.

### 3.4 Robustness: Sensitivity of the results to assumptions – optimistic and pessimistic scenarios

How do the results change if we assume there were some changes in expected mortality and/or registration completion during the pandemic, or if the mortality impact of the pandemic outside STAR12 differed from that in STAR12? In the optimistic scenario, as set out in Table 2, in STAR12 we obtain 1.6 excess deaths per 1K by May 2021. Nationally, we find 1.4–1.5 excess deaths per 1K by May 2021, rising to 2.0–2.1 by June. This equates to a total of 1.9–2.0M excess deaths up to May 2021, rising to 2.8M by June.

In the pessimistic scenario, as set out in Table 2, in STAR12 we obtain 2.7 excess deaths per 1K by May 2021. Nationally, we find 2.9–3.0 excess deaths per 1K by May 2021, rising to 3.9 by June. This equates to a total of 4.0–4.1M excess deaths up to May 2021, rising to 5.2M by June. The results are summarised in Table 3 below.

**Table 3:**
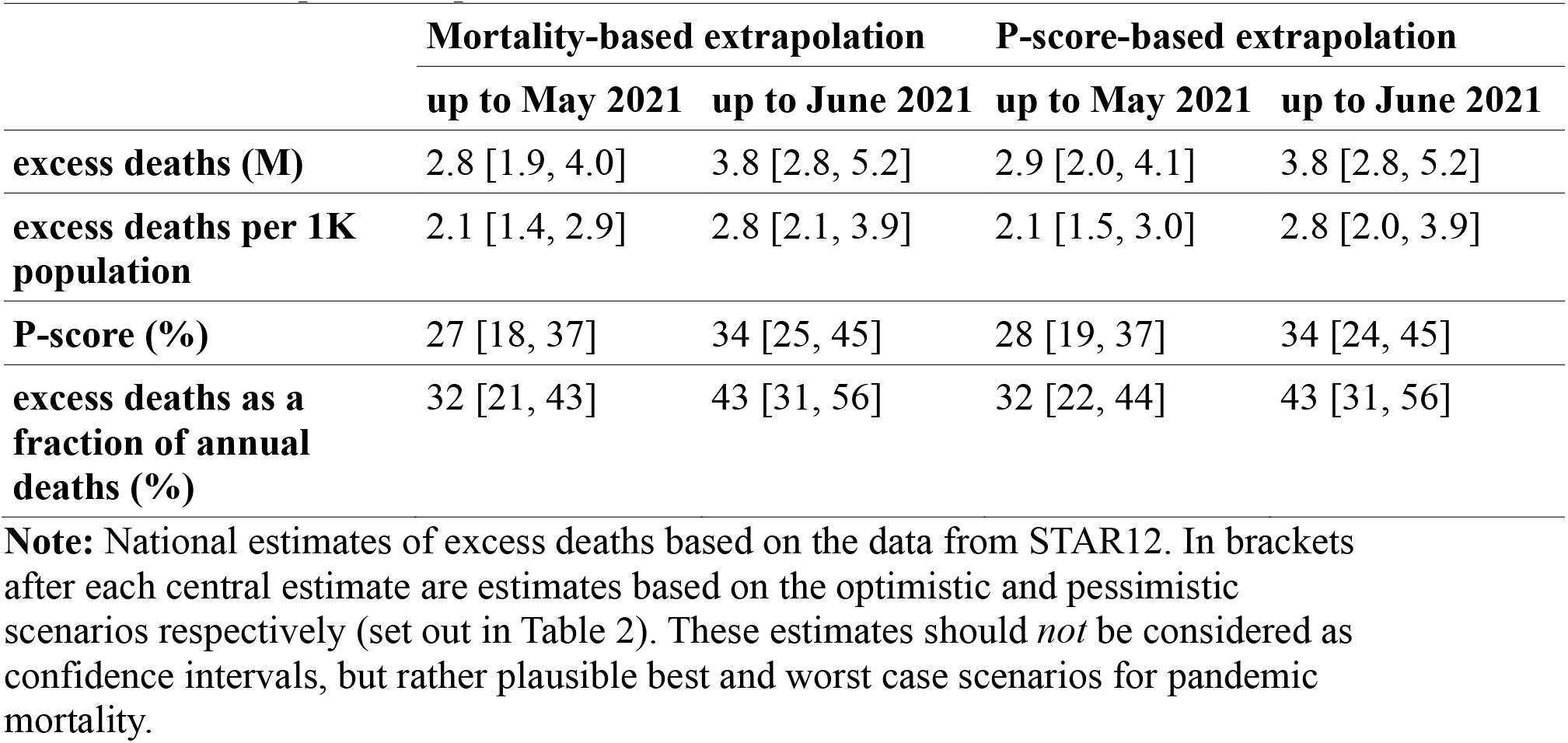
Central, optimistic, pessimistic estimates of excess deaths

|  | Mortality-based extrapolation |  | P-score-based extrapolation |  |
| --- | --- | --- | --- | --- |
|  | up to May 2021 | up to June 2021 | up to May 2021 | up to June 2021 |
| **excess deaths (M)** | 2.8 [1.9, 4.0] | 3.8 [2.8, 5.2] | 2.9 [2.0, 4.1] | 3.8 [2.8, 5.2] |
| **excess deaths per 1K population** | 2.1 [1.4, 2.9] | 2.8 [2.1, 3.9] | 2.1 [1.5, 3.0] | 2.8 [2.0, 3.9] |
| **P-score (%)** | 27 [18, 37] | 34 [25, 45] | 28 [19, 37] | 34 [24, 45] |
| **excess deaths as a fraction of annual deaths (%)** | 32 [21, 43] | 43 [31, 56] | 32 [22, 44] | 43 [31, 56] |

**Note:** National estimates of excess deaths based on the data from STAR12. In brackets after each central estimate are estimates based on the optimistic and pessimistic scenarios respectively (set out in Table 2). These estimates should *not* be considered as confidence intervals, but rather plausible best and worst case scenarios for pandemic mortality.

### 3.5 Robustness: 95% confidence intervals based on Monte-Carlo simulations

The optimistic and pessimistic scenarios presented above are based on the assumption of uniform changes – most importantly, uniform decreases or increases in registration coverage, across the states of STAR12. However, we can also examine how the estimates change if we allow independent variation in parameters associated with each state. To do this, we ran Monte-Carlo simulations, as described in section 2.3. We found that despite the fairly wide range of variation allowed in each state, 95% confidence intervals lie comfortably within the range from the optimistic and pessimistic scenarios.

In order to check robustness of these estimates to the choice of states we include in the analysis, we then carried out the same procedure with each of the 12 states omitted. Median estimates of excess deaths during April 2020–June 2021 varied between around 3.6M and 4.1M depending on the state omitted. The results of all the simulations are in Figure 2, and code used for the simulations is in replication materials (41). The results indicate that the central estimates, and the overall levels of uncertainty in the estimates, are not greatly affected by the omission of any individual state from the analysis.

**Figure 2:**
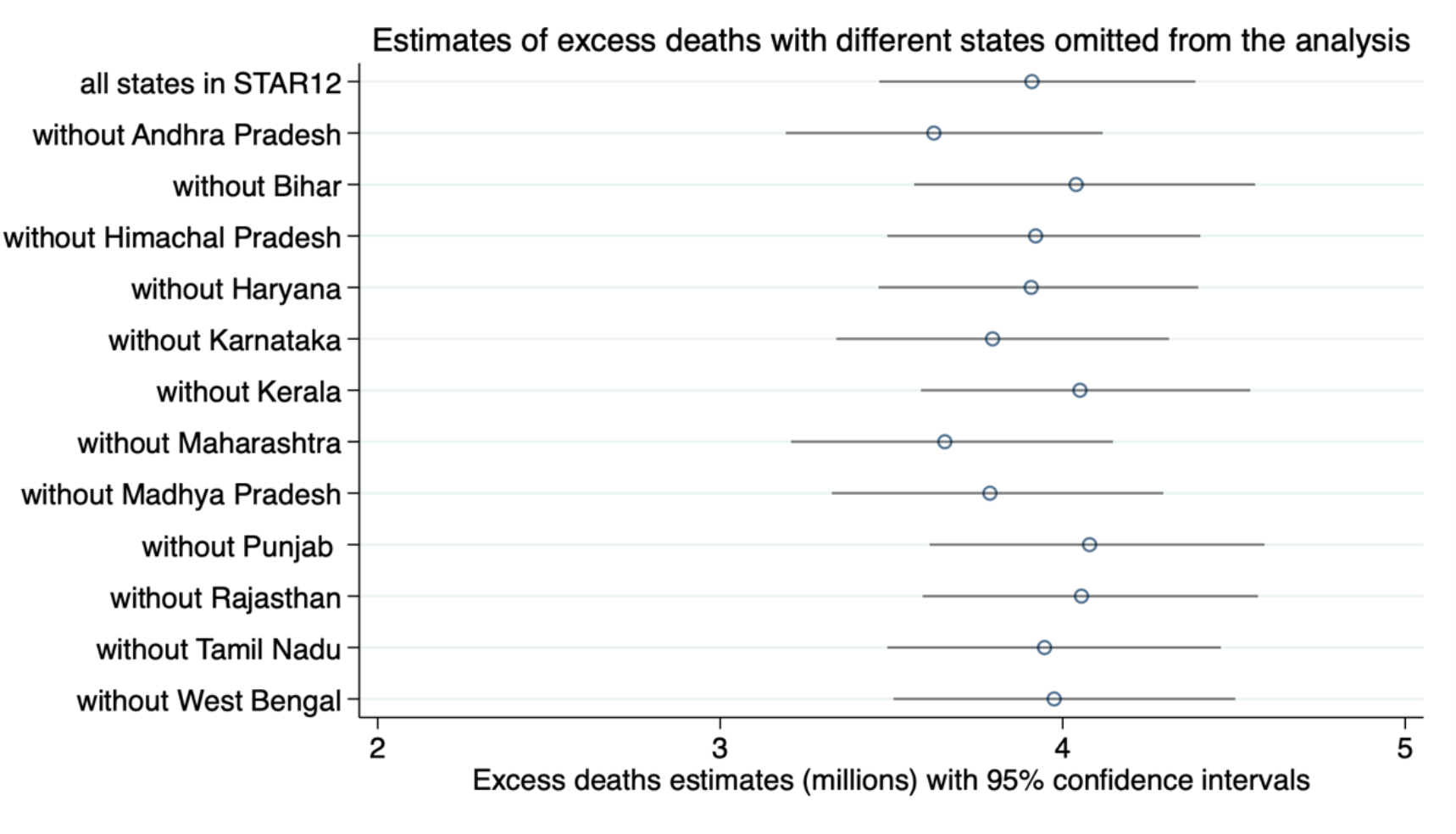
Estimates of excess deaths during April 2020–June 2021 and 95% confidence intervals, based on simulations

### 3.6 Robustness: Comparison of excess mortality with recorded COVID-19 deaths

In this section we assume no changes in expected deaths or registration completion, and use mortality-based extrapolations to get national estimates. The goal is to examine how excess mortality estimates align with official COVID-19 deaths over time.

We begin by noting the very high ratio of estimated excess deaths to official COVID-19 deaths. Overall, up to May 2021, estimated excess deaths relative to a 2019 baseline in STAR12 were 7.2 times recorded COVID-19 deaths in these states. The ratio was even higher nationally: during April 2020–May 2021 the estimated national excess death toll was 8.5 times the official COVID-19 toll. If we include estimates up to June 2021, this ratio rises to 9.5. In the optimistic scenario detailed in the previous Section this ratio is 6.9, while in the pessimistic scenario it is around 13.0.

The fact that the estimated ratio of excess deaths to recorded COVID-19 deaths nationally is higher than in STAR12 could reflect the fact that COVID-19 disease and death surveillance in the remaining states and territories was weaker than in STAR12. Recall that pre-pandemic registration completion was considerably higher in STAR12 than in remaining states. In addition, there is evidence that COVID-19 disease surveillance was weaker outside STAR12: by June 2021, per capita COVID-19 cases from STAR12 were about 70% higher than in the remainder of the country, even though seroprevalence data from the fourth national serosurvey (44,45) indicates that seroprevalence was similar across the strata. It is for this reason that we considered extrapolation based on official COVID-19 data to carry a higher risk of bias than extrapolation based on the level of excess mortality, or the P-score.

Qualitatively, national estimates of monthly excess deaths align well with COVID-19 deaths, as seen in the plot of the two data-sets in Figure 3. Note the very different scales.

**Figure 3:**
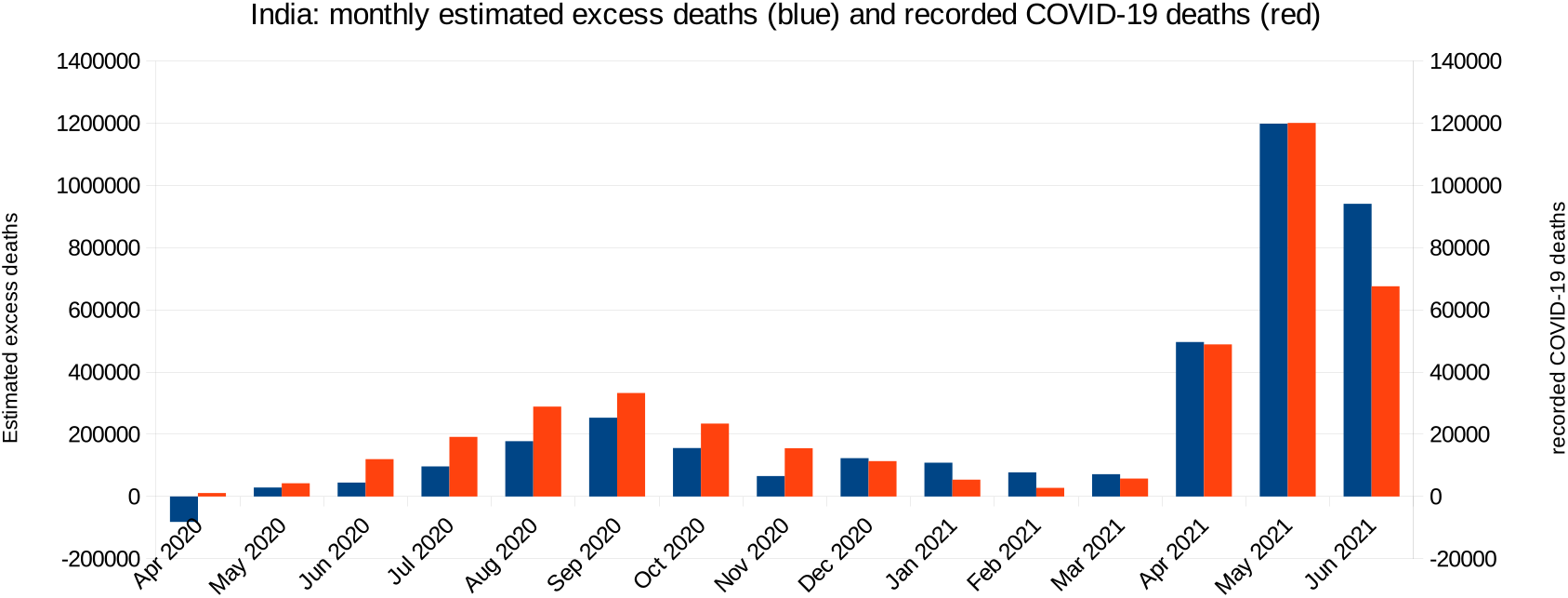
Estimated monthly excess deaths in India relative to a 2019 baseline, alongside recorded COVID-19 deaths. **Note:** The excess deaths are estimated from data in STAR12 according to methods described in the text and elaborated further in Appendix 2. Data on official COVID-19 deaths is from covid19india.org (COVID19India 2021)

During April 2020–Feb 2021, there is a strong linear association between estimated monthly excess deaths relative to a 2019 baseline and recorded COVID–19 deaths: the correlation coefficient is 0.84. During April 2020–May 2021, this rises to 0.98, and drops slightly to 0.96 over April 2020–June 2021. We note that this ecological analysis is associational, and is not intended to establish causality between excess deaths and recorded COVID-19 deaths. Instead, the fact that recorded COVID-19 deaths and excess deaths rise and fall together suggests that a large fraction of the excess deaths may in fact have been caused by COVID-19.

Comparisons between the time-course of excess deaths and COVID-19 deaths provide some clues as to how coverage in the data in STAR12 may have changed during the pandemic. The data is consistent with initial registration disruption, followed by recovery to 2019 levels or above. The question is how these effects summed over the duration of time for which we have data. March–April 2020 saw 10% fewer registrations in STAR12 than March–April 2019. Data from NFHS-5 indicates that the disruption may have caused an overall drop in registration completion during 2020 (22,24). On the other hand, January–March 2021 saw around 5% more death registrations than expected based on the ratio of excess to COVID-19 deaths in STAR12 during the remaining months of April 2020–May 2021. This could reflect improved coverage in the data by early 2021.

Given delays in registration, however, we should be cautious about interpreting changes in the ratio of excess deaths to COVID-19 deaths as indicative of changes in coverage. Karnataka, for example, saw 15% more death registrations during January–March 2021 than during the same period in 2019; this was despite the fact that coverage in the data was estimated to be 100% during 2019, and that during January–March 2021 official COVID-19 deaths in Karnataka were low. It is possible that delays in registration could account for the somewhat higher than expected registrations in states like Karnataka in the period between the country’s two COVID-19 waves.

#### Increasing under-ascertainment of COVID-19 deaths?

According to these estimates, the ratio of excess deaths to recorded COVID-19 deaths increased from around 6.7 during April 2020–February 2021, to around 11.3 during March–May 2021. This could reflect an increase in under-ascertainment of COVID-19 deaths during the huge 2021 surge or, possibly, an increase in non-COVID-19 excess deaths.

The scale of the increase in this ratio may be overestimated as a consequence of early disruption to registration and subsequent recovery. For example, during April 2020– February 2021 a modest 2% decrease in coverage in the data relative to 2019 levels leads to a 17% increase in estimated excess deaths during this period; in the other direction, a 5% increase in coverage in the data during March–May 2021 relative to 2019 levels, would reduce excess deaths estimates during this period by around 9%. These changes would have little effect on the overall estimate of pandemic excess deaths but would raise the first wave ratio of excess deaths to official COVID-19 deaths to 7.8, and lower the second–wave ratio to 10.3. Note that even given such a shift in registration coverage, the ratio would be higher during the second wave.

### 3.7 Robustness: Comparison of excess deaths and expected COVID-19 deaths based on international data

How do the estimates of excess mortality align with expectations of COVID-19 mortality given the scale of India’s epidemic? Based on India’s estimated 2021 age-structure (40), the meta-analysis of O’Driscoll et al. 2021 (46) predicts a COVID-19 infection fatality rate (IFR) of 0.25%, while the meta-analysis of Levin et al. 2020 (47) predicts COVID-19 IFRs of 0.42%–0.50% depending on the assumed age-distribution of the over-80s. There are some important caveats to such estimates: the meta-analyses differ, and both are based on 2020 COVID-19 fatality data primarily from high income countries and so, presumably, reflect the lethality of the original variants of SARS-CoV-2 circulating at the time, and the availability of healthcare in high-income settings.

Moreover, the predictions assume even spread of disease across different age groups. Having noted these caveats, what would IFR estimates of 0.25%–0.50% imply about COVID-19 deaths nationally, and how do these expectations align with our estimates of excess deaths?

#### Wave 1

The third national serosurvey carried out during December 2020–January 2021 estimated seroprevalence of 24.1% nationally, corresponding to approximately 325M infections nationwide (10). Allowing a variation in prevalence of 10% in either direction, IFR estimates of 0.25%–0.50% would imply 0.73–1.46M COVID-19 deaths. During April 2020–February 2021, our central estimate of 1.05M excess deaths lies comfortably within this range.

#### The whole pandemic period up to June 2021

Preliminary results from the fourth national serosurvey (45) found antibodies to SARS-CoV-2 in 62.3% of unvaccinated individuals sampled, corresponding to an estimated 839M COVID-19 infections by June 2021. Bearing in mind that these are unadjusted figures, and that some prior infections – especially older ones – may be missed, let us suppose that 750–1000M infections (equivalent to infection rates of 56% to 74%) had occurred by this point. IFR estimates of 0.25%–0.50% would then imply 1.9–5.0M COVID-19 deaths. Again, our central estimate of 3.8M excess deaths is comfortably within this range.

Thus India’s estimated excess mortality is broadly consistent with expected COVID-19 mortality based on meta-analyses using international data, and estimates of infection levels from seroprevalence surveys. In fact, the data are consistent with the following assertions, although none can be made definitively:

- The majority of excess deaths were likely COVID-19 deaths.
- A significant minority of excess deaths may have been either avoidable COVID-19 deaths (caused, for example, by unavailability of medical care or oxygen); or non-COVID-19 deaths caused, for example, by disruptions to health-care.
- During the second wave, more lethal variants and/or overwhelmed health systems may have driven up COVID-19 IFR despite increasing vaccination coverage. We need to treat this conclusion with some caution: registration disruption may have been particularly acute during national lockdown and the early part of the pandemic, leading to underestimation of first wave excess mortality. On the other hand, recovery of registration coverage to 2019 levels or higher during the second wave could lead to some overestimation of second wave excess mortality.

## 4. Discussion

The data from STAR12 allow us to infer with high confidence that STAR12 saw a major surge in mortality during the pandemic. The calculations here give estimates of 2.1 excess deaths per 1K population in these states during April 2020–May 2021. Repeated nationally, this level of excess mortality equates to around 2.8M excess deaths. Extending these estimates to June using more limited data we estimate around 3.8M excess deaths during April 2020–June 2021. Optimistic or pessimistic assumptions could shift these estimates by 20%–30% in either direction, while more up-to-date data are likely to push them up.

Given different age-structures and levels of development, comparing pandemic excess mortality in different countries is best done by considering excess deaths as a fraction of annual deaths. By this measure, India’s estimated excess death toll up to June 2021 was around 43% of its normal annual toll. Uncertainties in the data mean that this figure could plausibly lie between 31% and 56%. Using comparisons with international data on pandemic excess deaths as a percentage of annual deaths (34), even the lowest of these estimates places India amongst some of the hardest hit countries in the world.

The time-course of monthly excess deaths estimated using the process here displays a surprisingly strong association with recorded COVID-19 fatalities, rising and falling with the two waves of the epidemic. This suggests that the majority of these deaths reflect consequences of the pandemic, rather than underlying trends in mortality or death registration. This is not to say that these were all deaths from COVID-19: there may, indeed, have been some non-COVID-19 excess deaths. However, comparing the excess deaths estimates to expectations of COVID-19 mortality based on disease spread suggests that the majority of excess deaths were likely from COVID-19.

It is clear that official COVID-19 deaths have failed to capture the scale of pandemic excess mortality in India. If most excess deaths were, indeed, from COVID-19 then under-ascertainment of COVID-19 deaths has been high, with around 8–10 excess deaths for every recorded COVID-19 death. There is also evidence that under-ascertainment increased during the huge second wave.

There are several sources of uncertainty, which can lead to under- or over-estimation of excess mortality in STAR12 and nationally. These have already been discussed above, but we summarise and comment further on these below.

- *The data used is not up to date*. Delays in registration, and continued spread of disease, mean that we should expect further increases in estimates of excess deaths if more data becomes available. As some of the data we use is recorded by date of death (rather than date of registration), even totals for months where we already have data could rise.
- *The data for June 2021 is from only three states*. Estimates using this data carry greater uncertainty. It is possible that Andhra Pradesh, Karnataka and Punjab saw greater or later spread than the national average. Recall, however, that in these states June saw a sharp rise in the ratio of excess to COVID-19 deaths, indicative of registrations delayed during the massive surge in mortality in May 2021. We could see a similar pattern repeated in other parts of the country if June data becomes available.
- *There may be natural changes in yearly deaths on account of population growth and a changing CDR*. The likely scale of such changes was discussed in Section 2 and found to be fairly modest. SRS and UN estimates suggest a stable CDR in the years preceding the pandemic. Given an estimated population growth rate of around 1% a year, and a stable CDR, an increase in expected deaths of up to 2% for the pandemic period, relative to 2019 levels, is plausible. This would cause a reduction of about 7% in estimates of excess mortality.
- *National surge/excess mortality may not match the estimates from STAR12*. It is currently unclear in what direction data from more states and territories might push the estimates. But there is no convincing reason to believe the rest of the country was less hard hit than STAR12. Indeed, partial data (8,48,49) suggests that some of the absent states, such as Uttar Pradesh and Gujarat, have been very badly hit during the pandemic. The effects of these uncertainties are fairly easily quantified. For example, 20% lower or higher excess mortality in the remainder of the country relative to STAR12 changes national excess mortality by around 8%.
- *Death registration completion prior to the pandemic may have been overestimated*. We discussed earlier how death registration completion at the national level is likely overestimated in the 2019 CRS report, and even the sub-national estimates probably overestimate completion (22). Overestimating pre-pandemic completion is equivalent to underestimating pre-pandemic mortality. Taking into account some overestimation of pre-pandemic registration completion would push up estimates of total excess deaths in an easily quantifiable way. Excess deaths per million would go up, and so would the ratio of excess-to-COVID-19 deaths; but P-scores would not necessarily rise. Clearly, having accurate data on mortality is of importance even in non-pandemic times; the pandemic has, however, highlighted how critical it is.
- *Death registration levels could have changed during the pandemic*. Relatively small shifts in coverage in the data would significantly impact the estimates of excess mortality. For example, a 5% drop in coverage would increase excess mortality estimates by 20–25%. While registration coverage was gradually improving prior to the pandemic, there is evidence of considerable disruption to registration during the pandemic. Moreover, COVID-19 may have hit hardest in marginalised communities where death registration is weaker (50), lowering overall registration levels. The pandemic and associated lockdowns may also have changed the age-pattern of mortality, and if mortality increased most in ages where registration is poorer, this would again lower overall registration levels. Ultimately, reliable survey estimates will be needed to shed light on how registration coverage changed during the pandemic. However, the strong association between excess deaths and official COVID-19 deaths over time provides evidence that the bulk of the fluctuations in registered deaths reflect fluctuations in mortality driven by the pandemic, rather than trends in registration.

Ultimately, if more data becomes available it will be possible to unravel the potential biases with more confidence. But even the most optimistic and pessimistic views of the data do not change the story qualitatively. Consistent with the wide spread of disease, excess mortality has been high, and surveillance of COVID-19 deaths has been very weak.

## 5. Conclusion

Our findings highlight the extent to which populations are vulnerable to mortality crises in LMIC contexts and the extent to which disease surveillance systems under-estimate mortality. This points to the urgent need for robust systems to monitor all-cause mortality, as well as to improve the availability of real-time data from these systems. Our analysis reveals that data from India’s civil registration system can be challenging to analyse. Even so, this data provides key insights into India’s pandemic mortality crisis. We find that the surveillance of pandemic mortality in India has been extremely poor, with around 8–10 times as many excess deaths as officially recorded COVID-19 deaths. India is among the countries most severely impacted by the pandemic, and the pandemic is the gravest mortality crisis India has faced since its independence.

## Data Availability

All data used in the article is available at a single location on github

https://github.com/muradbanaji/IndiaACMdata

## Appendix 1. Estimates of crude death rate and registration completion in 2019

The 2019 report on Vital Statistics of India based on the Civil Registration System (1), provides both national and sub-national data on registered deaths, along with estimates of completion. The latter draw on estimates of the crude death rate (CDR) from the 2018 Sample Registration System annual statistical report (2). Recently, the 2019 Sample Registration System annual statistical report (3) has also been made available. Completion nationally or sub-nationally can be estimated by comparing total registered deaths to total expected deaths based on CDR and population estimates.

The <u>2018 SRS report</u> estimates India’s CDR at 6.2 per 1K, while this drops to 6.0 per 1K in the 2019 report. Using population projections (4), the SRS estimates of CDR imply 92–96% completion nationally in 2019. There are, however, several reasons to believe that the estimates of CDR at 6.0–6.2 could be too low, and hence the estimate of 92–96% completion too high.

### UN estimates

The United Nations estimated CDR in India to be 7.2 per 1K for the period 2015–20 (5), based on methodologies described by Gerland 2014 (6). This would imply an estimated registration completion during 2019 of only 80%.

### Combining sub-national estimates

We can combine sub-national estimates of registration completion in the 2019 CRS report to estimate national completion, and hence CDR. These calculations cap registration completion at 100%, i.e., if a region saw more deaths registered than expected from its estimated CDR, then registered deaths are taken as an estimate of total deaths. In each region we use the completion estimate to estimate total deaths, and summing these gives an estimated national death toll in 2019; from this we obtain national estimates of CDR and completion. This process gives an estimated national CDR in 2019 of 6.6 per 1K, and registration completion of around 86%. These estimates are based on subnational estimates of CDR from the 2018 SRS report. If, instead, we use data from the 2019 SRS report, then we obtain an estimated national CDR in 2019 of 6.5 per 1K, and registration completion of around 88%.

### Estimates based on age-stratified mortality rates

The 2018 and 2019 SRS reports give estimated death rates in different age groups. Projected population pyramids are available for 2016 and 2021, but not intervening years (4). Using the estimated age-specific mortality rates in the SRS reports, and the projected 2016 age distribution, we obtain an estimated national CDR of 6.6–6.8 per 1K, which would imply registration completion in 2019 of 84–87%. Using, instead, the 2021 age distribution, we obtain an estimated CDR of 7.3–7.5 per 1K, implying registration completion in 2019 of 76–79%. (Estimates depend on the fraction of the over-80s who are assumed to be over 85, which is not given in the projected population pyramids; we set this to be 0.375 as estimated in both SRS reports.)

### Estimates based on NFHS-5

NFHS-5 interviews were conducted between 2019 and 2021 (Government of India 2021) (7). The survey asked respondents about deaths of any usual family member in the previous few years, and whether the death was registered. From this data, we can compute a national registration level for 2018 of 73%, and hence a CDR for 2018 of 7.2. Assuming no change in the CDR between 2018 and 2019, we get estimated registration completion in 2019 of around 79%.

We thus find estimates of CDR nationally ranging from 6.0 to 7.5, corresponding to registration completion in 2019 ranging from 76% to 96%, with a cluster of estimates around 80%. The estimated completion of 86% derived from sub-national data in the 2019 CRS report may thus somewhat overestimate registration completion in India. We return to this point when discussing our results. More detail and full calculations are available at IndiaCOVIDmapping.org (8).

Figure A1 shows that for most states, levels of death registration completion observed in the NFHS-5 are lower than estimated official levels of death registration completion. The most likely explanation of over-estimation of official levels death registration completion is that the Crude Death Rate, estimated by the SRS, is under-estimated. When compared to official population projections and the 2011 Census, the SRS underestimates the proportion of people above age 65, and this contributes to lower Crude Death Rates in the SRS.

**Figure A1:**
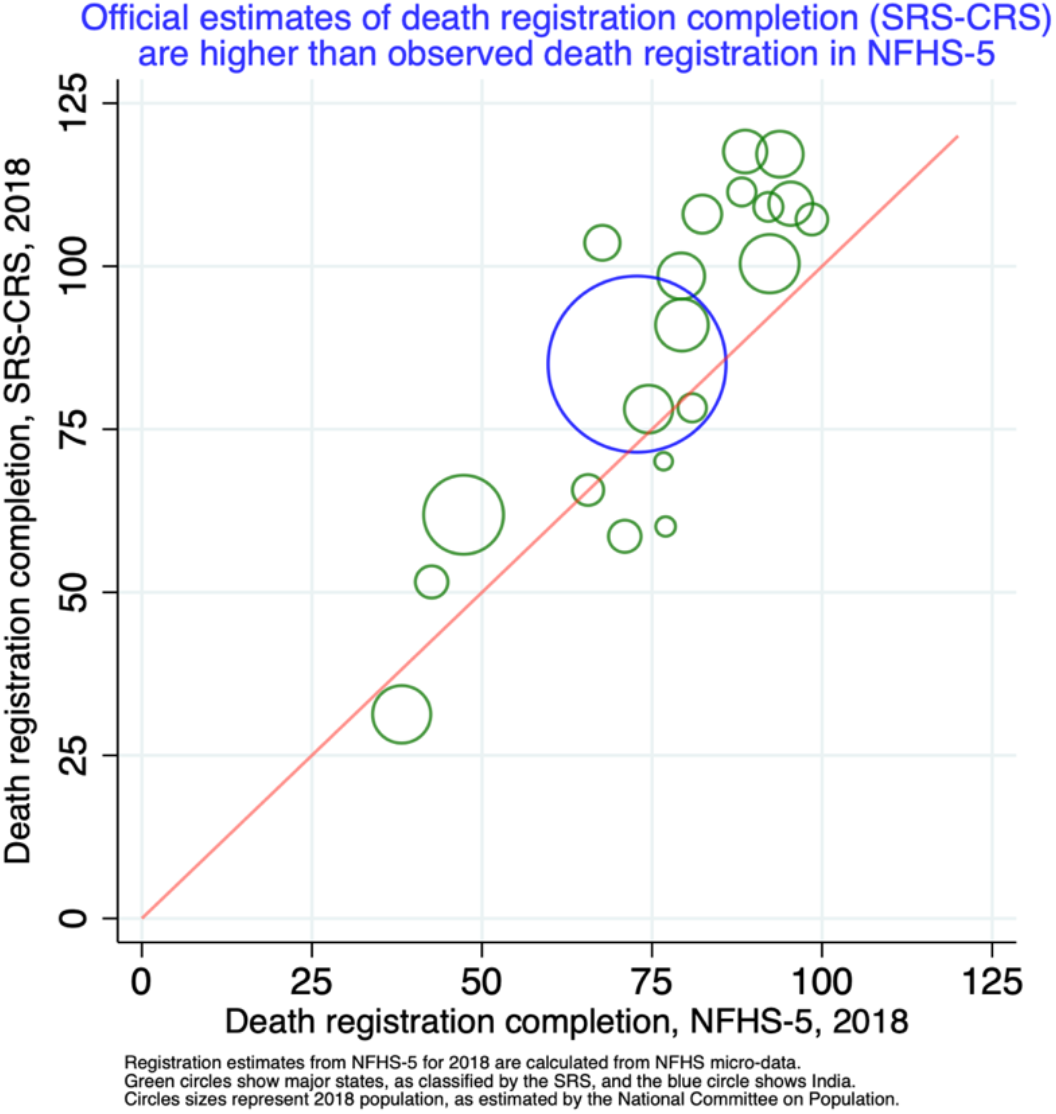
Comparison of estimates of death registration completion

## Appendix 2. Computing excess deaths, and sensitivity of the estimates to parameter changes

In order to estimate excess deaths in a given location over a given period, we need to estimate expected deaths and actual deaths. Given death registration data during some pandemic period and some comparable reference period, we additionally need estimates of:

1. coverage in the data during the reference period
2. coverage in the data during the pandemic periodic
3. expected changes in mortality between the reference period and pandemic period

With quantities in appropriate units, we can then compute:

1. (Pandemic deaths) = (pandemic registrations) / (pandemic coverage)
2. (Baseline deaths) = (baseline registrations) / (baseline coverage)
3. (Expected deaths) = (expected change) * (baseline deaths)
4. (Excess deaths) = (pandemic deaths) – (expected deaths)
5. P-score = 100*(excess deaths) / (expected deaths)

Finally, if we wish to extrapolate from some regions to others, we also need estimates of how the mortality impact may have varied between the regions for which we have data, and those for which we don’t.

We can examine how small changes in the key parameters affect our central estimates of excess deaths in STAR12 during April 2020–May 2021, and nationally. For each state, we make a small percentage change in each parameter of interest, namely baseline coverage in the data, baseline mortality, pandemic-period coverage in the data, and the difference in mortality impact in regions other than STAR12. The resulting outputs (excess deaths per capita and P-scores) are then compared with their baseline values.

Calculations are available online (9). From this process, we obtain the following estimates.

- *Changes in coverage*. A 1% (relative) decrease in pandemic period coverage in the data in each state relative to 2019 causes a 4.6% increase in excess deaths estimates in STAR12. Based on pre-pandemic trends and the disruption we see early in the pandemic, we might optimistically hope to see a relative increase in coverage in the data of 5% in states where this is possible; pessimistically, we might expect a relative decrease in coverage in the data of 5%. The reality is most likely that the competing effects of disruption and recovery summed in different ways in different states.
- *Changes in expected deaths*. A 1% increase in expected deaths during the pandemic period relative to 2019 (as a consequence of population growth and changes in CDR) would cause a 3.6% decrease in excess mortality estimates in STAR12. Based on pre-pandemic population growth and trends in CDR, we might expect year-on-year deaths to remain fixed or rise by at most 2%.
- *Errors in estimation of baseline mortality/coverage*. A 1% (relative) decrease in both baseline and pandemic period coverage in the data in each state causes a 1% increase in excess mortality estimates in STAR12. Optimistically, we might hope that the sub-national estimates of registration completion in the 2019 CRS report are accurate. A more pessimistic view would be that in some states they could have been overestimated by up to 15%.
- *Difference in mortality impact outside STAR12*. If regions not in STAR12 collectively saw a mortality impact 1% higher than in STAR12, this causes a 0.4% increase in the national mortality estimates. We consider it plausible that the mortality impact in STAR12 could differ by up to 20% from that in the remainder of the country, causing an 8% shift in our excess mortality estimates.

When we consider the effects and possible scale of the uncertainties in these parameters, it is clear that shifts in coverage in the data during the pandemic cause the greatest uncertainty in the estimates of excess mortality. By comparison, the likely effects of population growth, changes in CDR, and errors in estimates of pre-pandemic registration coverage are relatively small. There is also no compelling reason to believe that the mortality impact of the pandemic outside of STAR12 should have been considerably different from that in STAR12.

## References

1. Helleringer S, Queiroz BL. Measuring excess mortality due to the COVID-19 pandemic: progress and persistent challenges. International journal of epidemiology. 2021;

2. Knutson V, Aleshin-Guendel S, Karlinsky A, Msemburi W, Wakefield J. Estimating Global and Country-Specific Excess Mortality During the COVID-19 Pandemic. arXiv preprint 220509081. 2022;

3. Levin AT, Owusu-Boaitey N, Pugh S, Fosdick BK, Zwi AB, Malani A, et al. Assessing the burden of COVID-19 in developing countries: systematic review, meta-analysis and public policy implications. BMJ Global Health. 2022 May 1;7(5):e008477.

4. Lewnard JA, Mahmud A, Narayan T, Wahl B, Selvavinayagam TS, Laxminarayan R. All-cause mortality during the COVID-19 pandemic in Chennai, India: an observational study. The Lancet Infectious Diseases. 2022;22(4):463–72.

5. World Health Organization. Global Civil Registration and Vital Statistics: A Scaling Up Investment Plan 2015–2024. World Health Organization; 2014.

6. Gupta A. COVID-19 and the importance of improving civil registration in India. Center for the Advanced Study of India. 2020;

7. Karlinsky A. International completeness of death registration –2019. medRxiv. 2021;

8. Acosta RJ, Patnaik B, Buckee C, Balsari S, Mahmud Aß. All-cause excess mortality in the State of Gujarat, India, during the COVID-19 pandemic (March 2020-April 2021). medRxiv [Internet]. 2021; Available from: https://doi.org/10.1101/2021.08.22.21262432

9. Banaji M. Estimating COVID-19 infection fatality rate in Mumbai during 2020. medRxiv. 2021;

10. Murhekar MV, Bhatnagar T, Thangaraj JWV, Saravanakumar V, Kumar MS, Selvaraju S, et al. SARS-CoV-2 seroprevalence among the general population and healthcare workers in India, December 2020–January 2021. International Journal of Infectious Diseases. 2021;108:145–55.

11. Aburto JM, Schöley J, Kashnitsky I, Zhang L, Rahal C, Missov TI, et al. Quantifying impacts of the COVID-19 pandemic through life-expectancy losses: a population-level study of 29 countries. International journal of epidemiology. 2022;51(1):63–74.

12. Aburto JM, Kashyap R, Schöley J, Angus C, Ermisch J, Mills MC, et al. Estimating the burden of the COVID-19 pandemic on mortality, life expectancy and lifespan inequality in England and Wales: a population-level analysis. J Epidemiol Community Health. 2021;

13. Ramani S. On ‘excess deaths’ in Rajasthan. The Hindu [Internet]. 2021 Jul 4 [cited 2021 Sep 28]; Available from: https://www.thehindu.com/news/national/on-excess-deaths-in-rajasthan/article35136631.ece

14. Rukmini S. Madhya Pradesh saw nearly three times more deaths than normal after second wave of Covid-19 struck. Scroll.in [Internet]. 2021 [cited 2021 Sep 28]; Available from: https://scroll.in/article/996772/madhya-pradesh-saw-nearly-three-times-more-deaths-than-normal-after-second-wave-of-covid-19-struck

15. The Hindu Data Team. Data | India’s excess deaths could be highest among nations with the most recorded COVID-19 fatalities. The Hindu [Internet]. 2021 Jul 30 [cited 2021 Sep 28]; Available from: https://www.thehindu.com/data/data-indias-excess-deaths-could-be-highest-among-nations-with-most-recorded-covid-19-fatalities/article35636438.ece

16. Government of India. Vital Statistics of India Based on the Civil Registration System 2019. New Delhi: Office of the Registrar General of India, Ministry of Home Affairs, Government of India. 2021;

17. Banaji M, Gupta A. India: notes on estimates of crude death rate and registration coverage [Internet]. IndiaCOVIDMapping.org; 2022. Available from: https://www.indiacovidmapping.org/reports/mortality/IndiaRegistrationNotes.pdf

18. Rao C, Gupta A, Gupta M, Yadav AK. Premature adult mortality in India: what is the size of the matter? BMJ Global Health. 2021;6(6):e004451.

19. India COVID Mapping. Mapping the impact of the COVID-19 pandemic in India [Internet]. https://www.indiacovidmapping.org/. 2022. Available from: https://www.indiacovidmapping.org/

20. Rao C, John AJ, Yadav AK, Siraj M. Subnational mortality estimates for India in 2019: a baseline for evaluating excess deaths due to the COVID-19 pandemic. BMJ Glob Health. 2021 Nov 1;6(11):e007399.

21. International Institute for Population Sciences (IIPS), ICF. National Family Health Survey (NFHS-5), 2019-21:India. Mumbai: IIPS; 2022.

22. Banaji M, Gupta A, Paikra V. Mortality and Death Registration in India. The India Forum [Internet]. 2022 Aug 18 [cited 2022 Aug 30]; Available from: https://www.theindiaforum.in/article/mortality-and-death-registration-india

23. Rao C, Gupta M. The civil registration system is a potentially viable data source for reliable subnational mortality measurement in India. BMJ Global Health. 2020;5(8):e002586.

24. Rukmini S. A fact-check of every claim made by India to dispute WHO’s estimate of its Covid-19 deaths [Internet]. Scroll.in. https://scroll.in; 2022 x[cited 2022 Jun 10]. Available from: https://scroll.in/article/1024181/a-fact-check-of-every-claim-made-by-india-to-dispute-whos-estimates-of-covid-19-deaths

25. Hirschman C, Preston S, Loi VM. Vietnamese casualties during the American war: A new estimate. Population and Development Review. 1995;783–812.

26. Nepomuceno MR, Turra CM. The Population of Centenarians in Brazil: Historical Estimates from 1900 to 2000. Population and Development Review. 2020;46(4):813–33.

27. Zhang G, Zhao Z. Reexamining China’s fertility puzzle: Data collection and quality over the last two decades. Population and Development Review. 2006;32(2):293–321.

28. Anand A, Sandefur J, Subramanian A. Three new estimates of India’s all-cause excess mortality during the COVID-19 pandemic. Cent Glob Dev Work Pap. 2021;416.

29. Jha P, Deshmukh Y, Tumbe C, Suraweera W, Bhowmick A, Sharma S, et al. COVID mortality in India: National survey data and health facility deaths. Science. 2022;eabm5154.

30. Leffler CT, Lykins VJ, Das S, Yang E, Konda S. Preliminary Analysis of Excess Mortality in India During the COVID-19 Pandemic. The American Journal of Tropical Medicine and Hygiene. 2022;

31. Wang H, Paulson KR, Pease SA, Watson S, Comfort H, Zheng P, et al. Estimating excess mortality due to the COVID-19 pandemic: a systematic analysis of COVID-19-related mortality, 2020–21. The Lancet. 2022;399(10334):1513–36.

32. The Economist. The pandemic’s true death toll. The Economist [Internet]. 2021 Sep 27 [cited 2021 Sep 28]; Available from: https://www.economist.com/graphic-detail/2021/09/27/americas-pandemic-is-now-an-outlier-in-the-rich-world

33. Giattino C, Ritchie H, Roser M, Ortiz-Ospina E, Hasell J. Excess mortality during the Coronavirus pandemic (COVID-19). Our World in Data. 2020;

34. Karlinsky A, Kobak D. Tracking excess mortality across countries during the COVID-19 pandemic with the World Mortality Dataset. Elife. 2021;10:e69336.

35. ORGI. Sample registration system statistical report 2018. Office of the Registrar General and Census Commissioner of India, New Delhi: Government of India.; 2020.

36. ORGI. Sample registration system statistical report 2019. Office of the Registrar General and Census Commissioner of India, New Delhi: Government of India.; 2022.

37. United Nations, Department of Economic and Social Affairs, Population Division. World Population Prospects 2019 Data Booklet. ((ST/ESA/SER.A/424)).

38. ORGI. Sample registration system Bulletin (Various years) [Internet]. Office of the Registrar General and Census Commissioner of India, New Delhi: Government of India.; 2020. Available from: https://censusindia.gov.in/vital_statistics/SRS_Bulletins/

39. World Bank. Death rate, crude (per 1,000 people) - India | Data [Internet]. 2021 [cited 2021 Sep 28]. Available from: https://data.worldbank.org/indicator/SP.DYN.CDRT.IN?locations=IN

40. National Commission on Population. Population Projections for India and States 2011-2036. Report of the Technical Group on Population Projections Constituted by the National Commission on Population. 2019;

41. Banaji M. IndiaACMdata/National Estimates [Internet]. GitHub. 2022 [cited 2021 Sep 29]. Available from: https://github.com/muradbanaji/IndiaACMdata/tree/master/NationalEstimates

42. Government of India. Vital Statistics of India Based on the Civil Registration System 2020. New Delhi: Office of the Registrar General of India, Ministry of Home Affairs, Government of India. 2022;

43. Aron J, Muellbauer J. Transatlantic excess mortality comparisons in the pandemic. INET Oxford Working Paper No 2020-18. 2020;

44. PIB India. Centre advises States to conduct State-specific Sero Surveys in consultation with ICMR to generate district-level data on sero-prevalence [Internet]. 2021 [cited 2021 Sep 29]. Available from: https://pib.gov.in/pib.gov.in/Pressreleaseshare.aspx?PRID=1739902

45. PIB India. Implications of 4th Round of National Sero-Survey show that there is a ray of hope but there is no room for complacency. Non-essential travel must be discouraged and travel only if fully vaccinated - DG @ICMRDELHI #IndiaFightsCorona https://t.co/IwOs72LBAR [Internet]. @PIB_India. 2021 [cited 2021 Sep 28]. Available from: https://twitter.com/PIB_India/status/1417443573948043271

46. O’Driscoll M, Dos Santos GR, Wang L, Cummings DA, Azman AS, Paireau J, et al. Age-specific mortality and immunity patterns of SARS-CoV-2. Nature. 2021;590(7844):140–5.

47. Levin AT, Hanage WP, Owusu-Boaitey N, Cochran KB, Walsh SP, Meyerowitz-Katz G. Assessing the age specificity of infection fatality rates for COVID-19: systematic review, meta-analysis, and public policy implications. European journal of epidemiology. 2020;1–16.

48. Das S. Death Count In 24 UP Districts 43 Times More Than Official Covid-19 Toll — Article 14. Article 14 [Internet]. 2021 [cited 2021 Sep 27]; Available from: https://www.article-14.com/post/untitled-60cf605395758

49. Gupta A, Banaji M. The scale of Gujarat’s mortality crisis. The Hindu. 2021;

50. Bamezai A, Banaji M, Gupta A, Pandey S, Sharan MR, Sharma K, et al. Survey evidence of excess mortality in Bihar in the second COVID-19 surge. SocArxiv. 2021;

## References

1. Government of India. Vital Statistics of India Based on the Civil Registration System 2019. New Delhi: Office of the Registrar General of India, Ministry of Home Affairs, Government of India. 2021;

2. ORGI. Sample registration system statistical report 2018. Office of the Registrar General and Census Commissioner of India, New Delhi: Government of India.; 2020.

3. ORGI. Sample registration system statistical report 2019. Office of the Registrar General and Census Commissioner of India, New Delhi: Government of India.; 2022.

4. National Commission on Population. Population Projections for India and States 2011-2036. Report of the Technical Group on Population Projections Constituted by the National Commission on Population. 2019;

5. United Nations, Department of Economic and Social Affairs, Population Division. World Population Prospects 2019 Data Booklet. ((ST/ESA/SER.A/424)).

6. Gerland P. UN Population Division’s methodology in preparing base population for projections: case study for India. Asian Population Studies. 2014;10(3):274–303.

7. International Institute for Population Sciences (IIPS), ICF. National Family Health Survey (NFHS-5), 2019-21: India. Mumbai: IIPS; 2022.

8. India COVID Mapping. Mapping the impact of the COVID-19 pandemic in India [Internet]. https://www.indiacovidmapping.org/. 2022. Available from: https://www.indiacovidmapping.org/

9. Banaji M. IndiaACMdata/National Estimates [Internet]. GitHub. 2022 [cited 2021 Sep 29]. Available from: https://github.com/muradbanaji/IndiaACMdata/tree/master/NationalEstimates

